# Characteristic variables of acute ischemic stroke categorized according to body mass index classifications

**DOI:** 10.1101/2023.10.04.23296576

**Authors:** Joe Senda, Yoshinobu Amakusa, Tomoki Hirunagi, Ruido Ida, Hidenori Muro, Takahito Otani, Hiroko Nakagawa-Senda, Takehiro Naito, Takenori Kato, Toshinori Hasegawa, Katsuhiro Kawaguchi, Masahisa Katsuno

**Affiliations:** Department of Neurology, Komaki City Hospital, Komaki, Japan.; Department of Rehabilitation, Komaki City Hospital, Komaki, Japan.; Department of Neurology, Nagoya University Graduate School of Medicine, Nagoya, Japan.; Department of Rehabilitation Medicine, Nagoya City University Graduate School of Medical Sciences, Nagoya, Japan.; Department of Public Health, Nagoya City University Graduate School of Medical Sciences, Nagoya, Japan.; Department of Neurosurgery, Komaki City Hospital, Komaki, Japan.; Department of Cardiology, Komaki Cit y Hospital, Komaki, Japan.

**Author notes:** Address correspondence to: Joe Senda, MD, PhD., Department of Neurology and Rehabilitation, Komaki city hospital, 1-20 Jyoubushi Komaki 485-8520, Japan. Other authors’ E-mail address: Yoshinobu Amakusa, Tomoki Hirunagi, Takahito Otani, Hiroko Nakagawa-Senda, Ruido Ida, Hidenori Muro, Takehiro Naito, Takenori Kato, Toshinori Hasegawa, Katsuhiro Kawaguchi, Masahisa Katsuno.

**Keywords:** Body Mass Index, dyslipidemias, hyperlipidemias, ischemic stroke, overweight

## Abstract

**BACKGROUNND:** We investigated the characteristic variables of acute ischemic stroke categorized according to body mass index (BMI).

**METHODS:** From a registry of 1676 consecutive acute ischemic stroke cases, 965 cases (622 men and 434 women; mean age 71.53 ± 12.42 years) were selected based on eligibility criteria. These participants were divided into four cohorts according to BMI classifications: BMI < 18 (underweight), 18.5 ≤ BMI < 25 (normal weight), 25 ≤ BMI < 30 (overweight), and BMI ≥ 30 (obese). To avoid bias, propensity score analyses were performed; the confounding variables of sex and age were adjusted.

**RESULTS:** The underweight cohort (n = 82) had an older, largely female population. The overweight (n = 212) and obese cohorts (n = 48) had younger populations. The analyses revealed no significant differences in neurological outcomes among the four cohorts. Furthermore, higher risks of hyperlipidemia, dyslipidemia, and diabetes mellitus were associated with the higher BMI cohorts in participants with acute ischemic stroke. Findings revealed higher rates of usage of antidiabetic and antidyslipidemic medications in the overweight cohort (p = 0.007, p = 0.003; respectively) and raised values of triglycerides, low-density lipoprotein cholesterol, and glycated hemoglobin in the obese cohort (p = 0.044, p = 0.019, p < 0.001; respectively) than in the normal weight cohort. By contrast, the lower BMI cohort (underweight) did not have hyperlipidemia.

**CONCLUSIONS:** The importance of focusing on the management of dyslipidemia, hyperlipidemia, and hyperglycemia for acute ischemic stroke in patients who are overweight or obese has been highlighted.

## INTRODUCTION

Obesity is an important risk factor associated with a greater incidence of stroke ^1,2^. Body mass index (BMI) is widely used as an index of obesity derived from the variables of body weight and height. ^3^. According to a previous meta-analysis ^4^, the risk of stroke is positively correlated with BMI, and this correlation is stronger in men and patients with ischemic stroke. The findings of yet another study supported this meta-analysis, revealing that the BMI categories of overweight and obesity were associated with an increased risk only in men ^5^. It has been shown that there is a positive correlation between the whole range of BMI categories and ischemic stroke ^6^, with no evidence of a threshold or a J-shaped curve ^7,8^. Conversely, the BMI category of underweight in both sexes is also associated with a higher risk of ischemic stroke, and this category has a significantly higher rate of poor short and long-term outcomes than the BMI category of normal weight ^9,10,11^. Other studies regarding the association between BMI and the prognosis after acute ischemic stroke have shown that BMI has a non-linear association with 3-month outcomes in men with acute ischemic stroke ^8^, and that the BMI categories of underweight and obesity were generally associated with poorer outcomes ^12,13,14^.

The mechanisms of the nature in which BMI affects ischemic stroke are not fully understood. However, possible explanations include an elevated BMI increasing the risk of ischemic stroke by inducing hypertension, diabetes, dyslipidemia, and inflammation, which are known risk factors for atherosclerosis and thrombosis ^15,16^. These risk factors may impair cerebral blood flow and oxygen delivery, leading to ischemic injury ^16^. By contrast, a low BMI increases the risk of ischemic stroke through malnutrition, anemia, infection, and immunosuppression, which may compromise the brain’s resistance to ischemia. In addition, a low BMI may increase the risk of hemorrhagic transformation after ischemic stroke due to reduced clotting factors and platelets ^17,18^.

Some studies have shown that BMI exhibits an influence on the outcomes of ischemic stroke ^19,20^. However, the precise correlation of the wide-ranged BMI classification, including both categories of obesity and underweight, and characteristic variables of ischemic stroke have not been fully elucidated.

In the present study, we investigated the categorization of the characteristic variables and outcomes of the acute phase of ischemic stroke by BMI classifications in a single-center study. Many of these variables were determined by various analyses, such as blood tests and brain magnetic resonance imaging/angiography (MRI/MRA).

## METHODS

### Ethical Approval and Informed Consent

Informed consent was obtained from all study participants. This study was approved by the Ethics Committee of the Komaki City Hospital (approval number: 231003), and was performed in accordance with the provisions of the Declaration of Helsinki (as revised in Brazil 2013).

### Enrolment of Participants

We retrospectively registered 1676 consecutive cases of acute ischemic stroke that were hospitalized at the Komaki City Hospital from 2018 to 2022. As depicted in Figure 1, a total of 965 participants (622 men and 434 women, mean age 71.53 ± 12.42 years) were enrolled in the study. The inclusion criteria were as follows: (1) complete independence in performing ADLs, based on a score of 0 on the modified Rankin Scale in which the participant lived alone prior to the current episode of ischemic stroke ^21^; (2) no previous medical history of dementia, including Alzheimer’s disease; mild cognitive impairment or neurodegenerative diseases prior to the current ischemic stroke (this medical history was determined from the case files and interviews with participants and their families); and (3) participants who underwent brain MRI/MRA with the resultant diagnosis of the current ischemic stroke. As a result, the following cases were excluded: (i) in-hospital mortalities, due to missing data (n = 84); (ii) cases in which the participants had previous medical histories of both ischemic and hemorrhagic stroke (n = 395), dementia (n = 139), neurodegenerative diseases (n = 19); and (iii) cancer that was not in remission at the time of the current ischemic stroke (n = 20). In addition, participants who had incomplete independence in performing activities of daily living (ADLs) before the current ischemic stroke secondary to conditions such as orthopedic impediments (n = 20) were excluded. Finally, participants with missing data regarding their clinical examinations and investigations, such as laboratory blood tests and imaging findings, were not included in the statistical analysis (n = 34). The BMI, laboratory blood tests, and carotid ultrasounds were measured on admission, and the National Institutes of Health Stroke Scale (NIHSS) scores ^22^ were measured at discharge.

**Figure 1.**
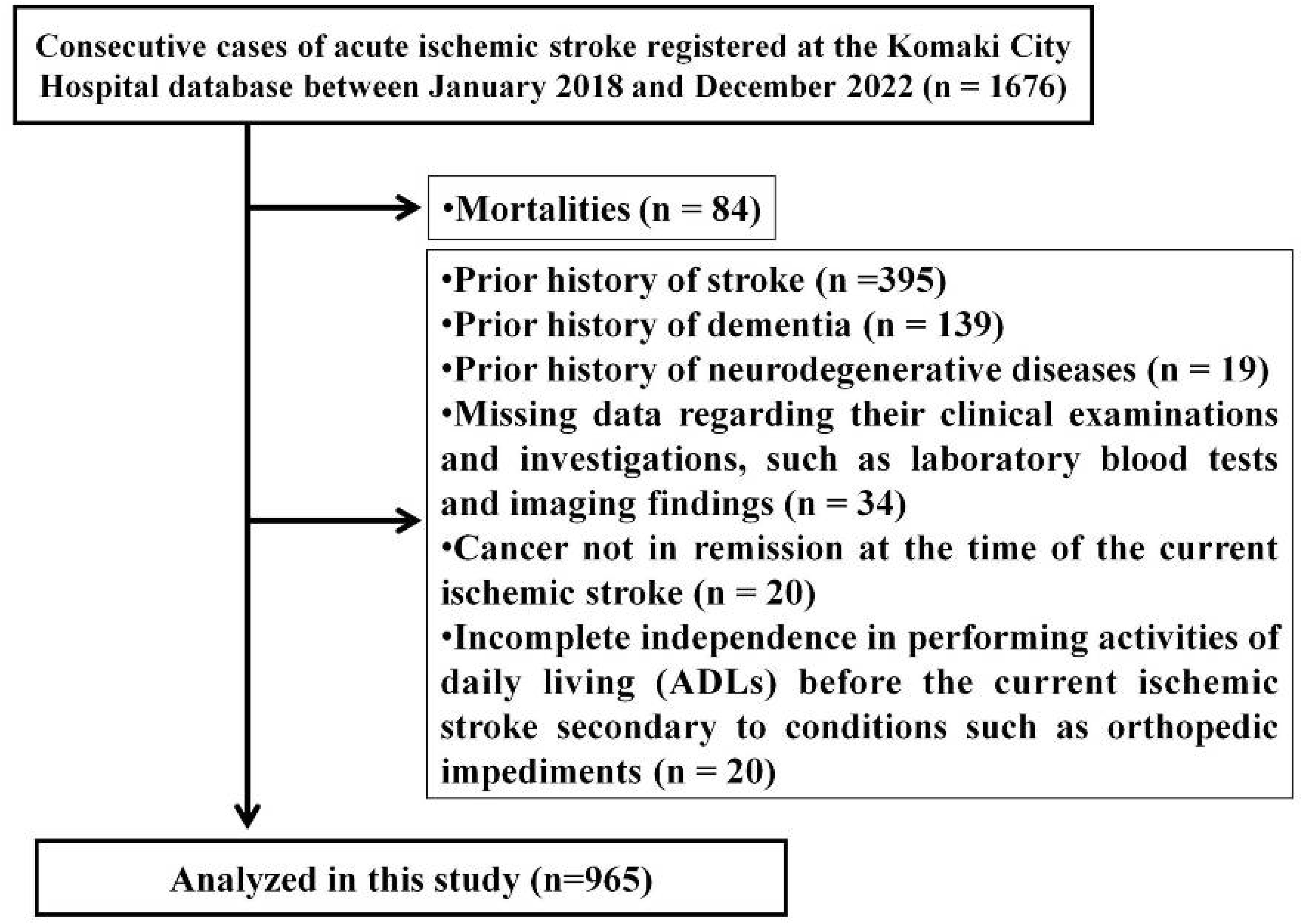
Flowchart Depicting Enrolment of Participants. Consecutive cases of acute ischemic stroke were registered at the Komaki City Hospital database between January 2018 and December 2022 (n = 1676). We excluded mortalities (n = 84). We also excluded participants who had previous medical histories of both ischemic and hemorrhagic stroke (n = 395), dementia (n = 139), neurodegenerative diseases (n = 19), and cancer that was not in remission at the time of the current ischemic stroke (n = 20). In addition, participants who had incomplete independence in performing activities of daily living (ADLs) before the current ischemic stroke secondary to conditions such as orthopedic impediments (n = 20) were excluded. Participants with missing data regarding their clinical examinations and investigations, such as laboratory blood tests and imaging findings, were not included in the statistical analysis (n = 34). Finally, 965 participants were included in the present study.

### Ischemic stroke subtype evaluation

In addition to the clinical categories of the National Institute of Neurological Disorders and Stroke-Classification of cerebrovascular diseases III of lacunar infarctions (LI), atherothrombotic (AT), and cardioembolic (CE) strokes ^23^, we appended artery-to-artery (A-to-A) embolic infarctions. Based on the Trial of ORG 10172 in Acute Stroke Treatment (TOAST) classification, A-to-A embolic infarctions that involve atheroembolic mechanisms can be distinguished from LI, CE, and AT. Embolic cases in which A-to-A embolic infarctions and CE strokes could not be diagnosed, or where the source of the embolism was unknown were classified as: “embolic stroke of undetermined source (ESUS).” Ischemic strokes secondary to a rare specific etiology, such as vasculitis or postoperative ischemic stroke, were classified as: “stroke of other determined etiology.” ^24^

### Analyses of MRI/MRA findings

Brain MRI/MRA were performed in all 965 participants on admission to the Komaki City Hospital. White matter lesions were classified using the Fazekas criteria for periventricular hyperintensities (PVH) and deep white matter hyperintensities (DWMH) using T2-weighted or fluid-attenuated inversion recovery images. PVH were graded from 0–3 as follows: Grade 0 = absence or rim only, Grade 1 = localized lesion depicted as pencil-thin lining or “caps”, Grade 2 = irregular hyperintensity with a smooth “halo”, and Grade 3 = irregular lesion extending into the deep white matter and periventricular region ^25^. DWMH were graded from 0–3 as follows: Grade 0 = absence, Grade 1 = punctate foci, Grade 2 = beginning confluence of foci, and Grade 3 = large confluent areas ^26^. On MRA, “stenosis positive” was defined as the presence of a stenosis or occlusion ≥ 50 % in the intracranial trunk arteries within the visible range ^27^.

### BMI categories

We categorized the participants into four cohorts: underweight (Cohort 1), normal weight (Cohort 2), overweight (Cohort 3), and obese (Cohort 4) defined as a BMI (kg/m^2^) of < 18.5, 18.5–24.9, 25.0–29.9, and ≥ 30, respectively ^28^.

### Statistical analyses

For the statistical analyses, continuous variables were presented as means ± standard deviations. Categorical variables were presented as percentages. A chi-square test was performed to compare categorical variables. A Steel-Dwass test was used for multiple comparisons. A Mann-Whitney U-test for each two-cohort comparison was performed to compare continuous variables. To avoid bias, confounding variables were balanced between cohorts, and propensity score analyses were conducted in which the variables of sex and age were adjusted for ^29^. In the propensity score analyses, one-to-one nearest neighbor matching without replacement was completed using a caliper width equal to 0.2 of the standard deviation of the propensity score. After the propensity score analyses, the comparisons among each of the four selected cohorts were conducted as follows: between (1) underweight (Cohort 1) vs. normal weight (Cohort 2), (2) normal weight (Cohort 2) vs. overweight (Cohort 3), (3) normal weight (Cohort 2) vs. obese (Cohort 4), and (4) overweight (Cohort 3) vs. obese (Cohort 4). For all statistical analyses, data were analyzed using EZR version 1.55 (Saitama Medical Center, Jichi Medical University, Saitama, Japan) ^30^, which is a graphical user interface for R (The R Foundation for Statistical Computing, version 4.1.2, Vienna, Austria; http://cran.r-project.org/). A p-value less than 0.05 was considered statistically significant.

## RESULTS

### Demographic characteristics of participants categorized according to BMI classification

The demographic characteristics of the study participants are summarized in Table 1. Comparisons of demographic data among the four cohorts revealed statistically significant differences in the variables of age and sex and other clinical indicators, such as hemoglobin values, leukoaraiosis grades, dyslipidemia, diabetes mellitus, and NIHSS scores. In particular, with age as a variable, the underweight cohort was statistically significantly older than the normal-weight cohort (p < 0.001); however, the overweight and obesity cohorts were statistically significantly younger than the normal weight cohort (p < 0.001). Regarding sex as a variable, the underweight cohort had a statistically significant larger number of women than that in the normal-weight cohort (p < 0.001). Furthermore, the underweight and obesity cohorts had statistically significantly severer NIHSS scores than those of the normal-weight cohort (p = 0.003, p = 0.021; respectively).

**Table 1:** Demographic characteristics of participants categorized according to body mass index classification.

|  | All (n=965) | Cohort 1 (n=82) | Cohort 2 (n=623) | Cohort 3 (n=212) | Cohort 4 (n=48) | p-value | p-value (post-hoc analysis as each two group comparison) |  |  |  |
| --- | --- | --- | --- | --- | --- | --- | --- | --- | --- | --- |
|  |  | BMI<18.5 | 18.5≤BMI<25 | 25≤BMI<30 | 30≤BMI | Steel-Dwass test | Cohort 1 vs 2 | Cohort 2 vs 3 | Cohort 2 vs 4 | Cohort 3 vs 4 |
| Age (years) | 71.53±12.42 | 78.91±9.93 | 72.81±11.42 | 68.42±12.31 | 55.95±13.39 | <0.001† | <0.001† | <0.001† | <0.001† | <0.001† |
| Male /Female | 622 / 343 | 37 / 45 | 413 / 210 | 140 / 72 | 32 / 16 | 0.002† | 0.001† | 0.999 | 0.999 | 0.999 |
| Body Mass Index (kg/m<sup>2</sup>) | 23.29±3.87 | 17.10±1.34 | 22.14±1.71 | 26.85±1.36 | 33.01±4.34 | <0.001† | <0.001† | <0.001† | <0.001† | <0.001† |
| Length of hospitalization (days) | 16.79±12.92 | 21.96±18.24 | 16.64±12.42 | 15.88±12.21 | 13.81±8.85 | 0.001† | 0.005† | 0.941 | 0.600 | 0.779 |
| Risk factor and comorbidity |  |  |  |  |  |  |  |  |  |  |
| Current smoker, n (%) | 218 (22.6%) | 10 (12.2%) | 135 (21.7%) | 53 (25.0%) | 20 (41.7%) | <0.001† | 0.107 | 0.007† | 0.999 | 0.321 |
| History of heart disease†, n (%) | 94 (9.7%) | 6 (7.3%) | 63 (10.1%) | 22 (10.4%) | 3 (6.3%) | 0.002† | 0.063 | 0.089 | 0.774 | 0.993 |
| Current medication |  |  |  |  |  |  |  |  |  |  |
| Hypertension, n (%) | 663 (68.7%) | 44 (53.7%) | 421 (67.6%) | 166 (78.3%) | 32 (66.7%) | 0.001† | 0.190 | 0.748 | 0.084 | 0.094 |
| Diabetes mellitus, n (%) | 240 (24.9%) | 13 (15.9%) | 139 (22.3%) | 66 (31.1%) | 22 (45.8%) | <0.001† | 0.554 | 0.049† | 0.001† | 0.211 |
| Dyslipidemia, n (%) | 445 (46.1%) | 25 (30.5%) | 280 (44.9%) | 115 (54.2%) | 25 (52.1%) | 0.704 | 0.854 | 0.995 | 0.823 | 0.818 |
| Ischemic stroke subtype |  |  |  |  |  |  |  |  |  |  |
| Lacunar infarction, n (%) | 125 (13.0%) | 2 (2.4%) | 75 (12.0%) | 38 (17.9%) | 10 (20.8%) | 0.001† | 0.043† | 0.133 | 0.290 | 0.966 |
| Atherothrombotic infarction, n (%) | 314 (32.5%) | 28 (34.1%) | 188 (30.2%) | 75 (35.4%) | 23 (47.9%) | 0.055 | 0.755 | 0.993 | 0.276 | 0.463 |
| Artery-to-artery embolic stroke, n (%) | 227 (23.5%) | 15 (18.3%) | 154 (24.7%) | 50 (23.6%) | 8 (16.7%) | 0.103 | 0.999 | 0.510 | 0.382 | 0.877 |
| Cardioembolic stroke, n (%) | 150 (15.5%) | 20 (24.4%) | 107 (17.2%) | 20 (9.4%) | 3 (6.3%) | 0.393 | 0.525 | 0.871 | 0.289 | 0.489 |
| Embolic stroke of undetermined source, n (%) | 120 (12.4%) | 15 (18.3%) | 80 (12.8%) | 23 (10.8%) | 2 (4.2%) | 0.002† | 0.575 | 0.987 | 0.591 | 0.727 |
| Stroke of other determined etiology, n (%) | 29 (3.0%) | 2 (2.4%) | 19 (3.0%) | 6 (2.8%) | 2 (4.2%) | 0.953 | 0.380 | 0.034 | 0.199 | 0.897 |
| Emergency endovascular treatment, n (%) | 90 (9.3%) | 12 (14.6%) | 58 (9.3%) | 9 (0.9%) | 1 (2.1%) | 0.125 | 0.429 | 0.999 | 0.322 | 0.372 |
| MRI-PVH grade | 0.699±0.802 | 1.012±0.936 | 0.717±0.786 | 0.613±0.779 | 0.312±0.657 | <0.001† | 0.032† | 0.218 | <0.001† | 0.028† |
| MRI-DWMH grade | 1.107±0.802 | 1.402±0.783 | 1.118±0.791 | 1.028±0.814 | 0.812±0.790 | <0.001† | 0.016† | 0.386 | 0.030† | 0.285 |
| MRA stenosis ≥50% or occlusion (+), n (%) | 296 (30.7%) | 31 (37.8%) | 186 (29.9%) | 65 (30.7%) | 14 (29.2%) | 0.531 | 0.459 | 0.996 | 0.999 | 0.997 |
| Echo ICA stenosis ≥30% (+), n (%) | 411 (42.6%) | 44 (53.7%) | 278 (44.6%) | 80 (37.7%) | 9 (18.8%) | <0.001† | 0.412 | 0.298 | 0.003† | 0.060 |
| Laboratory blood tests on admission |  |  |  |  |  |  |  |  |  |  |
| Hb (g/L) | 140.6±46.1 | 126.2±19.7 | 138.0±19.2 | 145.1±17.0 | 151.4±21.1 | <0.001† | <0.001† | <0.001† | <0.001† | 0.031† |
| BUN (mmol/L) | 5.018±2.757 | 7.097±3.671 | 6.347±2.590 | 5.930±2.853 | 5.426±2.198 | <0.001† | 0.761 | 0.001† | 0.005† | 0.446 |
| Cre (umol/L) | 80.21±75.41 | 77.70±76.47 | 87.69±64.27 | 78.23±25.72 | 75.23±25.72 | 0.006† | 0.009† | 0.826 | 0.881 | 0.699 |
| Total protein (g/dl) | 71.4±5.667 | 70.45±5.91 | 71.40±5.72 | 74.73±5.47 | 72.08±5.41 | 0.249 | 0.616 | 0.858 | 0.530 | 0.868 |
| Albumin (g/dl) | 40.07±4.35 | 37.91±4.84 | 39.84±4.34 | 41.37±3.58 | 40.91±4.97 | <0.001† | 0.003† | <0.001† | 0.311 | 0.982 |
| Total cholesterol (mmol/L) | 5.116±1.107 | 5.156±1.115 | 5.004±1.079 | 5.286±1.048 | 5.725±1.406 | <0.001† | 0.652 | 0.002† | 0.002† | 0.279 |
| Triglycerides (mmol/L) | 1.286±0.777 | 1.068±0.386 | 1.365±0.727 | 1.712±0.876 | 2.437±1.958 | <0.001† | 0.002† | <0.001† | <0.001† | 0.039† |
| HDL cholesterol (mmol/L) | 1.396±0.416 | 1.618±0.436 | 1.405±0.382 | 1.281±0.353 | 1.388±0.759 | <0.001† | <0.001† | <0.001† | 0.086 | 0.999 |
| LDL cholesterol (mmol/L) | 3.234±1.032 | 3.130±0.940 | 3.145±1.053 | 3.434±0.962 | 3.686±0.969 | <0.001† | 0.999 | <0.001† | 0.001† | 0.347 |
| HbA1C (%) | 6.583±3.020 | 5.960±0.783 | 6.573±3.571 | 6.648±1.624 | 7.475±1.981 | <0.001† | 0.088 | 0.044† | <0.001† | 0.003† |
| NIHSS at discharge (IQR/min-max) | 2.0 (1-7/0-37) | 5.5 (2-11/0-32) | 2.0 (1-7/0-35) | 2.0 (0-5/0-37) | 1.0 (1-3.25/0-17) | <0.002† | 0.003† | 0.723 | 0.021† | 0.090 |
| Nasogastric tubefeeding (+), n (%) | 66 (6.8%) | 9 (11.0%) | 49 (7.9%) | 7 (3.3%) | 1 (2.1%) | 0.048† | 0.908 | 0.114 | 0.473 | 0.971 |

MRI: magnetic resonance imaging, PVH: periventricular hyperintensity, DWMH: deep white matter hyperintensity, MRA: magnetic resonance angiography, ICA: internal carotid artery, Hb: hemoglobin, BUN: blood urea nitrogen, Cre: creatinine, HDL: high density lipoprotein, LDL: low density lipoprotein, HbA1c: glycohemoglobin, NIHSS: national institute of health stroke scale, IQR: interquartile range. Dagger indicates statistically significant differences.

### Characteristics of the participants analyzed with propensity score matching methods

A propensity score analysis was conducted to avoid bias due to the confounding variables of age and sex. Table 2 depicts the characteristics of the participants as variables analyzed with propensity score matching methods. Table 2 (a) reveals the results of the underweight (Cohort 1) and normal weight (Cohort 2) cohorts after the propensity score analyses. These results revealed that the values of the triglycerides (TG, p = 0.013) and glycated hemoglobin (HbA1C, p = 0.002) in Cohort 1 were statistically significantly lower than those in Cohort 2; however, the value of high-density lipoprotein cholesterol (HDL-C) in Cohort 1 was statistically significantly higher than that in Cohort 2 (p = 0.003). Similarly, Table 2 (b) reveals the results of the normal weight (Cohort 2) and overweight (Cohort 3) cohorts and showed that the rates of the use of antihypertensive (p < 0.001), antidiabetic (p = 0.007), and antidyslipidemic (p = 0.003) medications in Cohort 3 were statistically significantly higher than those in Cohort 2. In addition, the values of hemoglobin (Hb) (p< 0.001), albumin (p= 0.001), TG (p< 0.001), and low-density lipoprotein cholesterol (LDL-C) (p= 0.004) in Cohort 3 were statistically significantly higher than those in Cohort 2. Furthermore, the value of HDL-C (p< 0.001) in Cohort 3 was statistically significantly lower than that in Cohort 2. Table 2 (c) reveals the results of the normal weight (Cohort 2) and obese (Cohort 4) cohorts. These results showed that the rates of the use of antidiabetic (p < 0.001) and disease type as atheroembolic (p = 0.025), as well as the values of Hb (p = 0.043), TG (p = 0.044), LDL-C (p = 0.019), and HBA1C (p < 0.001) in Cohort 4 were all statistically significantly higher than those in Cohort 2. Table 2 (d) reveals the results of the overweight (Cohort 3) and obese (Cohort 4) cohorts. There were no statistically significant differences between the values of the clinical indicators for these two cohorts. Additionally, regarding the NIHSS scores, the propensity score analyses revealed no statistically significant differences among the four cohorts.

**Table 2:** Characteristics of the participants analyzed with propensity score matching methods.

|  | (a) underweight vs normal weight |  |  | (b) normal weight vs overweight |  |  | (c) normal weight vs obese |  |  | (d) overweight weight vs obese |  |  |
| --- | --- | --- | --- | --- | --- | --- | --- | --- | --- | --- | --- | --- |
|  | Cohort 1 (n=82) | Cohort 2 (n=82) | p-value | Cohort 2 (n=209) | Cohort 3 (n=209) | p-value | Cohort 2 (n=42) | Cohort 4 (n=42) | p-value | Cohort 3 (n=42) | Cohort 4 (n=42) | p-value |
|  | BMI<18.5 | 18.5≤BMI<25 |  | 18.5≤BMI<25 | 25≤BMI<30 |  | 18.5≤BMI<25 | 30≤BMI |  | 25≤BMI<30 | 30≤BMI |  |
| Age (years) | 78.91±9.93 | 78.44±8.80 | 0.746 | 68.89±12.15 | 68.69±12.2 | 0.866 | 58.07±12.45 | 58.36±12.51 | 0.917 | 58.60±11.98 | 58.76±11.84 | 0.949 |
| Male /Female | 37 / 45 | 34 / 48 | 0.639 | 132 / 77 | 140 / 69 | 0.413 | 33 / 9 | 29 / 13 | 0.327 | 30 / 12 | 28 / 14 | 0.489 |
| Body Mass Index (kg/m<sup>2</sup>) | 17.10±1.34 | 21.95±1.77 | <0.001† | 22.02±1.79 | 26.85±1.36 | <0.001† | 22.69±1.57 | 33.05±4.59 | <0.001† | 26.85±1.37 | 33.17±4.59 | <0.001† |
| Length of hospitalization (days) | 21.96±18.24 | 17.05±12.48 | 0.054 | 15.26±10.32 | 15.94±12.28 | 0.543 | 11.98±8.16 | 14.02±9.24 | 0.285 | 14.508±9.35 | 14.21±9.36 | 0.889 |
| Risk factor and comorbidity |  |  |  |  |  |  |  |  |  |  |  |  |
| Current smoker, n (%) | 10 (12.2%) | 8 (9.8%) | 0.620 | 64 (30.6%) | 52 (24.9%) | 0.191 | 17 (40.5%) | 17 (40.5%) | 1.000 | 14 (33.3%) | 16 (42.9%) | 0.375 |
| History of heart disease†, n (%) | 6 (7.3%) | 12 (14.6%) | 0.136 | 23 (11.0%) | 22 (10.5%) | 0.875 | 3 (7.1%) | 3 (7.1%) | 1.000 | 4 (9.5%) | 3 (7.1%) | 0.697 |
| Current medication |  |  |  |  |  |  |  |  |  |  |  |  |
| Hypertension, n (%) | 44 (53.7%) | 60 (73.2%) | 0.009† | 133 (63.6%) | 163 (78.0%) | 0.001† | 21 (50.0%) | 29 (69.0%) | 0.077 | 32 (76.2%) | 29 (69.0%) | 0.469 |
| Diabetes mellitus, n (%) | 13 (15.9%) | 24 (29.3%) | 0.040† | 41 (19.6%) | 65 (31.1%) | 0.007† | 4 (9.5%) | 18 (42.9%) | <0.001† | 13 (31.0%) | 20 (47.6%) | 0.121 |
| Dyslipidemia, n (%) | 25 (30.5%) | 36 (44.0%) | 0.076 | 84 (40.2%) | 114 (54.5%) | 0.003† | 16 (38.1%) | 23 (54.8%) | 0.129 | 27 (64.3%) | 22 (52.4%) | 0.274 |
| Ischemic stroke subtype |  |  |  |  |  |  |  |  |  |  |  |  |
| Lacunar infarction, n (%) | 2 (2.4%) | 8 (9.8%) | 0.051 | 25 (12.0%) | 38 (18.2%) | 0.076 | 6 (14.3%) | 10 (23.9%) | 0.272 | 5 (11.9%) | 9 (21.4%) | 0.247 |
| Atherothrombotic infarction, n (%) | 28 (34.1%) | 22 (26.8%) | 0.251 | 67 (32.1%) | 73 (34.9%) | 0.535 | 11 (26.2%) | 21 (50.0%) | 0.025† | 13 (31.0%) | 23 (54.8%) | 0.028† |
| Artery-to-artery embolic stroke, n (%) | 15 (18.3%) | 15 (18.3%) | 1.000 | 47 (22.5%) | 50 (23.9%) | 0.729 | 8 (19.0%) | 6 (14.3%) | 0.564 | 11 (26.2%) | 6 (14.2%) | 0.179 |
| Cardioembolic stroke, n (%) | 20 (24.4%) | 20 (24.4%) | 1.000 | 31 (14.8%) | 20 (9.6%) | 0.101 | 7 (16.7%) | 2 (4.7%) | 0.079 | 5 (11.9%) | 2 (4.8%) | 0.241 |
| Embolic stroke of undetermined source, n (%) | 15 (18.3%) | 14 (17.1%) | 0.839 | 28 (13.4%) | 23 (11.0%) | 0.456 | 7 (16.7%) | 1 (2.4%) | 0.026† | 4 (9.5%) | 1 (2.4%) | 0.171 |
| Stroke of other determined etiology, n (%) | 2 (2.4%) | 3 (3.7%) | 0.652 | 11 (5.2%) | 5 (2.4%) | 0.127 | 3 (7.1%) | 2 (4.7%) | 0.649 | 4 (9.5%) | 1 (2.4%) | 0.171 |
| Emergency endovascular treatment, n (%) | 12 (14.6%) | 8 (9.8%) | 0.343 | 22 (10.5%) | 19 (9.1%) | 0.623 | 4 (9.5%) | 1 (2.4%) | 0.171 | 7 (16.7%) | 1 (2.4%) | 0.026† |
| MRI-PVH grade | 1.012±0.936 | 0.829±0.587 | 0.173 | 0.622±0.655 | 0.617±0.673 | 0.950 | 0.357±0.544 | 0.357±0.527 | 1.000 | 0.405±0.504 | 0.333±0.508 | 0.620 |
| MRI-DWMH grade | 1.402±0.783 | 1.207±0.556 | 0.098 | 1.001±0.550 | 1.029±0.588 | 0.807 | 0.738±0.633 | 0.881±0.545 | 0.397 | 0.905±0.646 | 0.881±0.857 | 0.895 |
| MRA stenosis ≥50% or occlusion (+), n (%) | 31 (37.8%) | 25 (30.5%) | 0.326 | 53 (25.3%) | 65 (31.1%) | 0.193 | 11 (26.2%) | 11 (26.2%) | 1.000 | 17 (40.5%) | 10 (23.9%) | 0.104 |
| Echo ICA stenosis ≥30% (+), n (%) | 44 (53.7%) | 42 (51.2%) | 0.756 | 86 (41.1%) | 79 (37.8%) | 0.485 | 13 (31.0%) | 7 (16.7%) | 0.127 | 12 (28.6%) | 7 (16.7%) | 0.197 |
| Laboratory blood tests on admission |  |  |  |  |  |  |  |  |  |  |  |  |
| Hb (g/L) | 126.2±19.7 | 131.6±16.1 | 0.058 | 138.2±19.9 | 145.3±16.9 | <0.001† | 142.7±17.0 | 151.5±21.8 | 0.043† | 150.0±13.2 | 148.8±20.9 | 0.751 |
| BUN (mmol/L) | 7.097±3.671 | 6.844±2.899 | 0.625 | 16.49±6.54 | 16.67±7.99 | 0.796 | 4.787±1.674 | 5.401±2.185 | 0.153 | 5.766±1.867 | 5.476±1.553 | 0.529 |
| Cre (umol/L) | 77.70±76.47 | 83.10±61.88 | 0.603 | 76.91±53.92 | 78.68±25.64 | 0.611 | 69.84±22.10 | 75.14±25.63 | 0.312 | 77.79±21.22 | 76.91±27.40 | 0.861 |
| Total protein (g/dl) | 70.45±5.91 | 71.40±5.72 | 0.648 | 71.14±4.42 | 71.53±4.31 | 0.480 | 71.31±4.47 | 72.38±4.13 | 0.360 | 71.98±3.64 | 71.90±4.49 | 0.950 |
| Albumin (g/dl) | 37.91±4.84 | 39.84±4.34 | 0.156 | 40.00±3.42 | 41.38±2.83 | 0.001† | 41.21±2.66 | 41.21±3.74 | 1.000 | 42.12±2.33 | 40.36±3.88 | 0.053 |
| Total cholesterol (mmol/L) | 5.156±1.115 | 4.873±1.185 | 0.115 | 5.083±1.141 | 5.276±10.51 | 0.061 | 5.232±0.922 | 5.651±1.222 | 0.080 | 5.496±1.044 | 5.756±1.434 | 0.344 |
| Triglycerides (mmol/L) | 1.068±0.386 | 1.194±0.872 | 0.013† | 1.355±0.386 | 1.724±0.766 | <0.001† | 1.666±1.001 | 2.217±1.252 | 0.044† | 1.841±0.963 | 2.398±1.750 | 0.108 |
| HDL cholesterol (mmol/L) | 1.618±0.436 | 1.424±0.385 | 0.003† | 1.444±0.436 | 1.275±0.349 | <0.001† | 1.460±0.402 | 1.416±0.805 | 0.752 | 1.190±0.330 | 1.425±0.800 | 0.082 |
| LDL cholesterol (mmol/L) | 3.130±0.940 | 3.009±1.628 | 0.441 | 3.156±0.941 | 3.426±0.966 | 0.004† | 3.186±0.856 | 3.653±0.935 | 0.019† | 3.664±0.969 | 3.690±0.975 | 0.904 |
| HbA1C (%) | 5.960±0.783 | 6.573±3.571 | 0.002† | 6.652±1.494 | 6.644±1.096 | 0.984 | 5.819±0.430 | 7.452±1.380 | <0.001† | 6.880±2.040 | 7.480±1.940 | 0.169 |
| NIHSS at discharge (IQR/min-max) | 5.5 (2-11/0-32) | 3.0 (1-7/0-35) | 0.234 | 2.0 (1-5/0-33) | 2.0 (1-4/0-37) | 0.473 | 2.0 (1-3/0-21) | 1.0 (1-2/0-17) | 0.313 | 2.0 (1-3/0-21) | 1.0 (1-2/0-17) | 0.313 |
| Nasogastric tubefeeding (+), n (%) | 9 (11.0%) | 9 (11.0%) | 1.000 | 13 (6.2%) | 13 (6.2%) | 1.000 | 2 (7.1%) | 1 (2.4%) | 0.311 | 0 (0%) | 1 (2.4%) | 0.320 |

## DISCUSSION

In the present study, each of the adverse outcomes (as determined by the NIHSS scores) of acute ischemic stroke in the underweight and obese cohorts were statistically significantly severer than those in the normal cohort. However, a propensity score analysis inclusive of the variables of age and sex revealed no statistically significant difference in these adverse outcomes among the four cohorts; thus, we hypothesized that these adverse outcomes were strongly affected by the variables of age and sex ^31^.

In addition, we found statistically significant differences in hyperlipidemia and dyslipidemia among the BMI cohorts. Previous literature reporting on the correlations among BMI, hyperlipidemia, and ischemic stroke were based on cohort studies ^27,32,33,34^, and there is little information regarding investigations performed at the acute phase of ischemic stroke. The findings of the current study revealed that participants with acute ischemic stroke in the overweight and obese cohorts had severe hyperlipidemia. These cohorts had decreased values of HDL-C and increased values of LDL-C and TG; in particular, the values of LDL-C and TG were severely increased in the obese cohort. In addition, the elevated percentage values of HBA1C in the obese cohort were indicative of diabetes mellitus. Previous studies have shown that patients with a high BMI also have metabolic risk factors, including hyperlipidemia and diabetes mellitus ^35,36,37^; therefore, we postulate that these metabolic risk factors result in ischemic stroke occurring in relatively younger patients.

By contrast, the underweight cohort did not have increased values of LDL-C and TG and only had a statistically significantly increased HDL-C compared to the normal weight cohort. The sarcopenia risk for stroke-associated infections is associated with a larger BMI category of ≥ 18.5; thus, we hypothesize that the pathophysiological mechanisms of ischemic stroke are different for our cohorts with larger BMIs ^15,16,38^. In addition, it is a well-known fact that a larger body fat distribution, as indicated by an elevated BMI, has a close relationship with the occurrence of metabolic disorders ^39^.

The present study has several limitations. First, this was not a population-based study; patients were referred to this single acute care hospital that has a stroke unit. Second, we could not further explore the data as per the stroke subtypes. Recent reports have revealed that the overweight and obese BMI categories are inversely correlated with poor outcomes of the cardioembolic and small vessel disease subtypes; however, there is no statistically significant relationship between stroke outcomes and BMI classification for the large artery disease subtypes ^40^. Furthermore, to firmly ratify conclusions, it must be considered that these findings were observed in a relatively small population size. Therefore, further cohort studies with larger population sizes are needed. Finally, this study may have been influenced by unknown variables.

In conclusion, the findings of this current study revealed that higher risks of hyperlipidemia, dyslipidemia, and diabetes mellitus were associated with the higher BMI categories, viz., the overweight and obese cohorts, in participants with acute ischemic stroke. This was evidenced by the higher rates of usage of antidiabetic and antidyslipidemic medication in the overweight cohort, and the raised values of TG, LDL-C, and HBA1C in the obese cohort than those in the normal weight cohort. By contrast, in participants with acute ischemic stroke, the lower BMI category, viz., the underweight cohort, did not have hyperlipidemia. Thus, our findings suggest that different pathophysiological mechanisms of ischemic stroke may exist for higher BMIs. Furthermore, the importance of focusing on the management of dyslipidemia, hyperlipidemia, and hyperglycemia for the acute phase of ischemic stroke in patients who are overweight or obese has been highlighted. The management and control of acute ischemic stroke related to BMI provides potentially useful information to encourage further studies regarding the relationships between BMI, metabolic risk factors, and the incidence of acute ischemic stroke.

## Data Availability

All data underlying the results are available as part of the article and no additional source data are required, and the raw data that support the findings of this study are available from the corresponding author, Joe Senda, upon reasonable request.

## Acknowledgments

We thank the staff of the Komaki City Hospital for collecting the data used in this study.

